# Interactions Between Gut Microbiome, Genetic Risk, and Obesity: Insights into Metabolic Pathways and Host-Microbe Dynamics

**DOI:** 10.1101/2025.06.18.25329842

**Authors:** Shreyas V. Kumbhare, Inti Pedroso, Karthik M. Muthukumar, Santosh K. Saravanan, Carmel Irudayanathan, Garima Sharma, Ranjan Sinha, Daniel E. Almonacid

## Abstract

**Background:** Obesity is a multifactorial metabolic disorder driven by genetic predisposition and gut microbiome composition, influencing key metabolic pathways through the gut-brain axis, gut-liver interactions, immune modulation, and insulin resistance. This study integrates gut microbiome and genetic data to uncover microbial and genetic signatures linked to obesity across these metabolic dimensions.

**Methods:** We analyzed gut microbiome composition and polygenic risk scores (PGS) for BMI in a cohort of 3,319 individuals, with genetic data available for 2,883 participants. Regression analyses were performed to identify bacterial taxa and microbial metabolic pathways associated with baseline BMI. Interaction analyses assessed how genetic predisposition modifies microbiome associations with BMI. A subset of individuals (n = 180) with longitudinal microbiome data was analyzed to examine microbial changes associated with BMI reduction.

**Results:** Several bacterial taxa, including Oscillospiraceae, *Faecalibacterium*, and Christensenellaceae, were inversely associated with BMI, while *Lachnoclostridium* and *Dorea* exhibited positive associations. Microbial pathways related to neurotransmitter metabolism, amino acid degradation, and short-chain fatty acid synthesis were significantly associated with BMI, highlighting their potential role in obesity. Interaction analyses revealed that BMI-PGS attenuated the association of butyrate synthesis and tryptophan degradation with BMI, while *Faecalibacterium* exhibited a stronger association in individuals with high genetic risk. Longitudinal analysis showed that shifts in microbial composition accompanied BMI reduction, particularly increases in Christensenellaceae and reductions in *Dorea*.

**Conclusions:** The gut microbiome is intricately linked to obesity through its modulation of metabolic pathways across the gut-brain axis, gut-liver axis, inflammation, and insulin resistance. Genetic predisposition further modifies these relationships, emphasizing the need for personalized approaches in microbiome-based obesity interventions.

## Introduction

Obesity is a complex, multifactorial condition influenced by genetic, environmental, and lifestyle factors. It is closely associated with several metabolic and inflammatory disorders, including type 2 diabetes, cardiovascular diseases, and metabolic-associated fatty liver disease (MAFLD) [1]. Increasing evidence highlights the critical role of the gut-brain axis, gut-liver axis, inflammation and immunity, and insulin resistance in the pathophysiology of obesity, revealing intricate interconnections between metabolic, neurological, and immune functions [2].

The gut microbiome is crucial in regulating these four metabolic dimensions, influencing host metabolism, inflammation, and immune homeostasis. The gut-brain axis is mediated through microbial metabolites such as short-chain fatty acids (SCFAs), neurotransmitters, and gut-derived peptides that influence appetite regulation and neuroinflammation [3]. Similarly, the gut-liver axis is impacted by gut microbial dysbiosis, which alters bile acid metabolism, increases intestinal permeability, and promotes hepatic steatosis [4]. Chronic low-grade inflammation, driven by gut microbiota-mediated immune activation, is a key contributor to insulin resistance and metabolic dysfunction [5]. Additionally, genetic predisposition plays a role in shaping the gut microbiota composition, which in turn modulates host metabolic responses [6]. Understanding the interplay between genetics, gut microbiota, and metabolic health is essential for developing targeted therapeutic strategies to modulate the microbiome to mitigate obesity and its associated metabolic disorders.

Recent studies have identified specific gut microbial pathways in obesity through these four dimensions. For instance, gut-brain axis alterations have been linked to dysregulated tryptophan metabolism, which affects serotonin availability and contributes to appetite dysregulation [7,8]. Similarly, the gut-liver axis is modulated by small molecular carboxylic acids, which act as mediators between gut microbiota and hepatic inflammation in obesity [9]. Chronic low-grade inflammation in obesity has been associated with increased lipopolysaccharide (LPS) production, leading to hepatic and systemic immune activation [2]. Furthermore, insulin resistance is exacerbated by gut microbial short-chain fatty acids (SCFAs) and bile acid metabolism, which influence glucose homeostasis and peripheral insulin sensitivity [10].

Given the complexity of obesity, understanding its pathophysiology through these four metabolic dimensions rather than as a singular disorder is imperative. Traditional obesity treatments focus on caloric restriction and physical activity, yet emerging evidence suggests that personalized interventions targeting the gut microbiome could provide more effective and sustainable solutions [11]. A multidimensional approach that integrates gut-brain signaling, gut-liver metabolism, immune modulation, and insulin resistance mechanisms will be crucial in developing precision medicine for obesity management [4].

This study approaches obesity biomarker discovery through a multidimensional lens, integrating the gut-brain axis, gut-liver interactions, immune modulation, and insulin resistance. Rather than viewing the gut microbiome and genetics as isolated factors, we examine their interplay across these metabolic dimensions to uncover microbial and genetic signatures contributing to obesity risk and progression. By incorporating PGS, we further assess how genetic predisposition shapes gut microbiome associations with BMI.

## Methods

### Study design and participants

Participants in this study were enrolled in the Digbi Health personalized care program for weight management (Digbi Control™), a commercially available program. All participants were ≥18 years of age, resided in the United States at the time of the study, and had access to the program through their employer-sponsored benefits.

This observational study included individuals ≥18 years old as a mandatory inclusion criterion. Exclusion criteria comprised individuals who self-reported antibiotic use at enrollment and those without weight data available near the sampling time point.

The Institutional Review Board of E&I Review Services reviewed and approved the study protocol (protocol code #18053, approved 05/22/2018). Before being included in the study, all participants provided electronic informed consent as part of their written consent process.

### Intervention and data collection

The Digbi Health intervention program is a digital therapeutics platform designed to support weight loss and address weight-related inflammatory disorders and associated comorbidities. The program integrates genetic and gut microbiome profiling, baseline participant information, and continuous health monitoring through an AI-powered mobile application. Upon enrollment, participants received online access to the Digbi Health app and were required to complete a comprehensive health questionnaire. They were also provided with a Bluetooth-compatible digital weighing scale, buccal swab, and stool sampling kits, which were shipped directly to them for the self-collection of biological samples.

The Digbi Health app facilitated continuous tracking of participants’ weight, dietary intake (via uploaded food photographs), and other lifestyle metrics. Health coaches assigned meals a nutrient density score based on their inflammatory potential, fiber diversity, and expected insulin response. The details of the personalized intervention are described elsewhere [12]. Individual level metadata: demographics and health outcomes used in the analyses are provided in Table S1.

### Biological sample collection and processing

Subjects self-collected fecal samples using Mawi Technologies iSWAB Microbiome collection kit (Model No. ISWAB-MBF-1200) and saliva samples using buccal swabs, adhering to standardized protocols. Fecal DNA extraction was performed with the Qiagen MagAttract Power Microbiome DNA Kit on an automated liquid-handling DNA extraction system. The extracted DNA underwent 16S rRNA gene V3–V4 region amplification and sequencing on the Illumina MiSeq platform (2 × 300 bp paired-end sequencing) at Akesogen Laboratories, Atlanta, GA. Microbiome data processing included sequence demultiplexing, denoising, and amplicon sequence variant (ASV) generation using DADA2 in QIIME2 (version 2021.4) [13]. Genotyping analysis was conducted using Affymetrix Direct-to-Consumer Array version 2.0 on the GeneTitan platform at Akesogen Laboratories. Genotype calling and data analysis followed best practices established in prior research [14].

### Genetic and gut microbiome analysis

Genotype call files were obtained from the genotyping facility and formatted in VCF format, with quality control steps including discordant genotype removal and left normalization. Beagle v5.3 [15,16] was used for phasing and imputation with the 1KG Project as the reference panel [17], retaining variants with imputation r² ≥ 0.8 for downstream analyses. PGS for obesity, diabetes, and related conditions were computed as described elsewhere (Table S2) [18]. PGS was calculated using PLINK2 (“plink2 --score”) [19], with genetic scores coded such that higher values indicate greater inherited predisposition.

For microbiome data, initial quality control included removing primers, low-quality bases, and non-bacterial ASVs (e.g., Euryarchaeota, Chloroplast, Mitochondria) or unassigned taxa at the phylum level. Taxa were agglomerated at the genus level, reducing the sparsity of the abundance matrix to 49.29% and removing singletons. The abundance matrix was rarefied to 30,000 reads per sample (minimum sample depth) with 500 iterations in QIIME2, resulting in 183 genera in total. Functional microbial pathway abundances related to gut and neuroactive metabolites were computed as described elsewhere [20,21]. All raw abundances were centered-log ratio (CLR) transformed unless stated otherwise.

### Statistical analysis

We used PERMANOVA to perform community-level (microbiome) multivariate association with variables based on the abundance matrix using the Bray-Curtis dissimilarity and using the “vegan” package (“adonis2” function) in R [22]. Principal Component Analysis (PCA) was performed using robust CLR (rCLR) transformed microbiome counts. To identify taxa and functional pathways associated with baseline BMI (T1), we utilized linear models implemented on the GAMLSS software package for R [23]. We used the following regression formulas to test the association with BMI: “abundance ∼ 1 + Gender + Age + BMI” and with the following options ‘gamlss(. , control=gamlss.control(c.crit = 0.001, n.cyc = 2000), family = BEZI())’, where “abundance” corresponds to the rarified counts of the taxa or functions divided by the total counts of the sample, “BMI” is the BMI at the time of sample collection, and “BEZI” is the beta distribution family (Beta Zero Inflated). We performed corrections for multiple hypothesis tests using the local FDR (lfdr) methodology implemented on the “ashr” software package for R [24,25], using the beta and standard error obtained in the regression analysis and with the option “mixcompdist=’normal’.” We set a local ‘lfdr’ 0.05 as the cutoff to declare statistical significance. To identify PGS associated with baseline BMI, we used a similar regression model with “family = NO” (normal distribution family), where PGS were z-score transformed using standard deviation. Principal components (PC1–PC5) of ancestry scores were included as covariates to account for population stratification and minimize confounding due to genetic ancestry differences. Additionally, we examined interactions between microbiome taxa or functional pathways and PGS to assess potential gene-microbiome influences on BMI, recognizing that host genetics may modulate microbial composition and metabolic function. For the longitudinal subset analysis, the BMI percent change was calculated as [(BMI at T1-BMI at T2) / (BMI at T1)] * 100, while the change in taxa or pathway abundances was determined by calculating the absolute change, T1: baseline and T2: follow-up during the intervention.

## Results

### Cohort characteristics

The study cohort consisted of a total of 3,319 individuals with gut microbiome data, out of which 2,883 individuals had genetic data. The median age of the participants was 49 years (SD: 12.24). In terms of gender distribution, the cohort was predominantly female (n = 2,619, 78.9%), with a smaller proportion of male participants (n = 700, 21.1%). The median baseline BMI was 32.16 (SD: 7.64), with participants categorized into different BMI groups: non-obese (n = 285, 8.6%), overweight (n = 889, 26.8%), Class I obesity (n = 1,030, 31.0%), Class II obesity (n = 583, 17.6%), and Class III obesity (n = 532, 16.0%).

### Gut microbial composition at baseline

The gut microbiome composition at baseline was analyzed using PCA (Figures 1A and 1B), showing a continuous gradient rather than distinct clustering across BMI-based obesity classes. Several bacterial taxa, including *Ruminococcus*, *Prevotella*, *Flavonifractor*, and members of the Ruminococcaceae family, were identified as key contributors to the variation in microbial composition.

**Figure 1:**
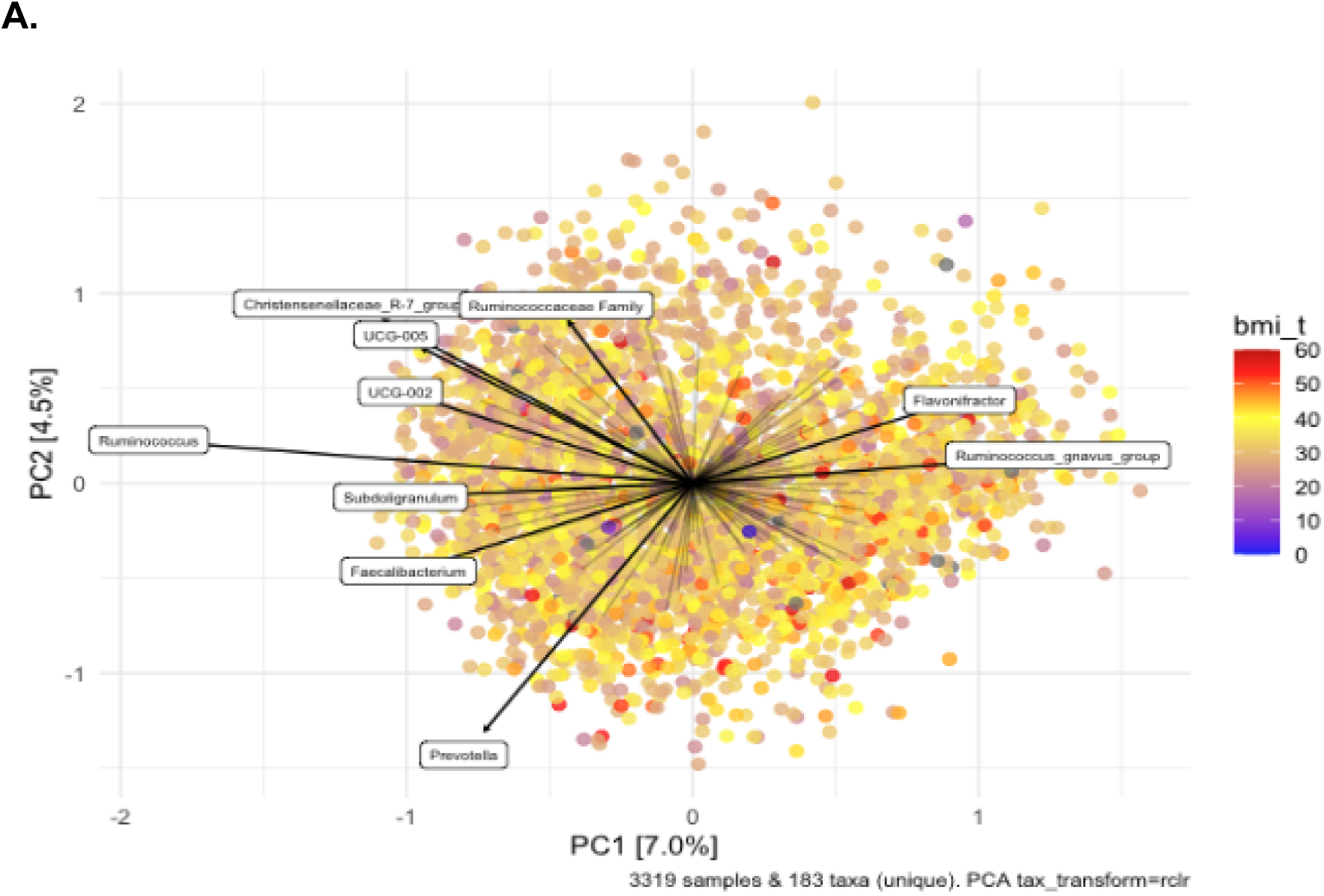
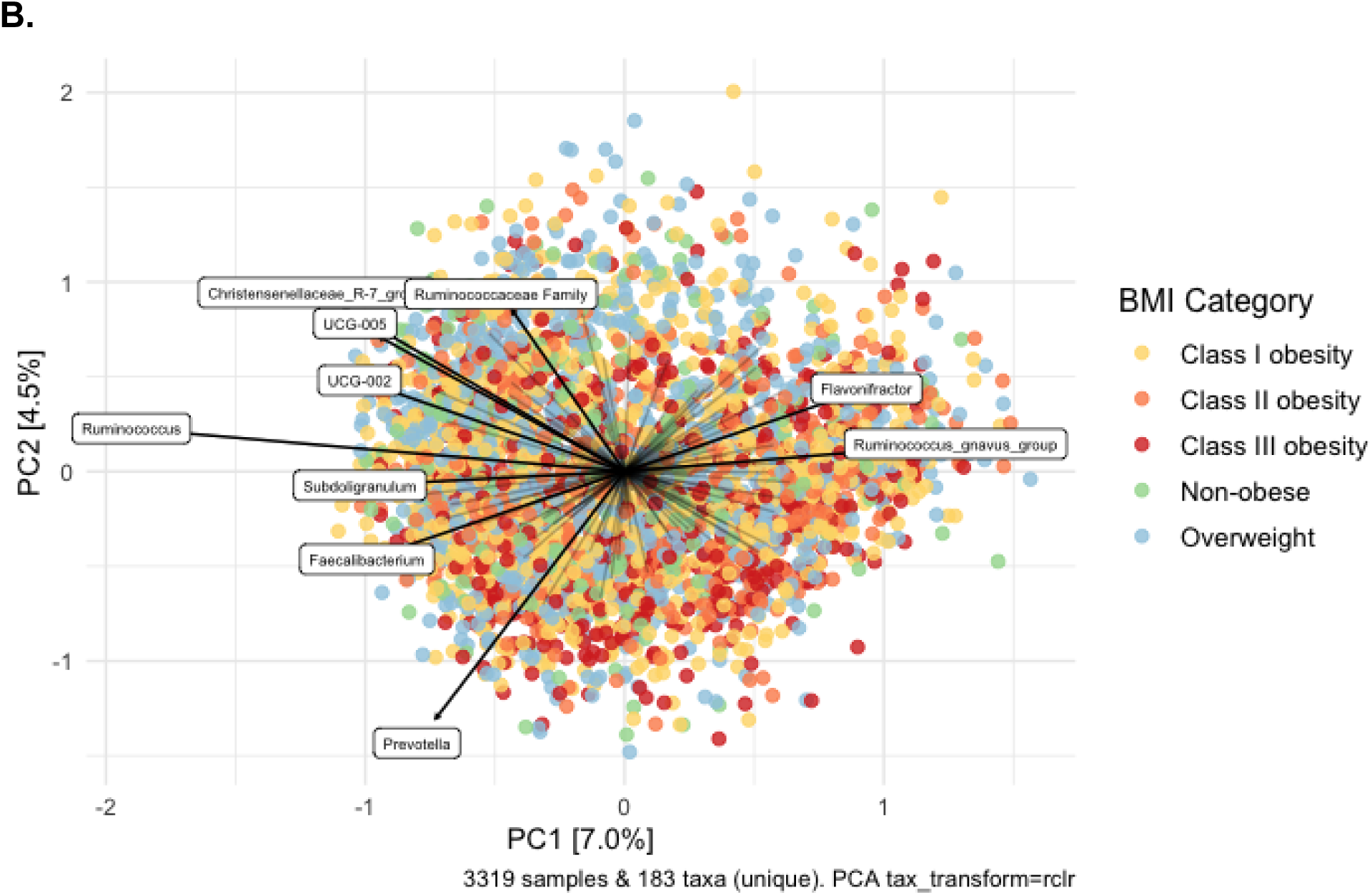
Principal Component Analysis (PCA) of Gut Microbiome Composition at Baseline. PCA plot displaying the gut microbiome composition of 3,319 individuals at baseline, with samples color-coded based on BMI as a (A) continuous variable and (B) BMI levels/categories (non-obese, overweight, and Class I–III obesity). The first two principal components (PC1 and PC2) explain 7.0% and 4.5% of the variance, respectively. Bacterial taxa annotated with arrows indicate key contributors to microbiome variation.

To evaluate the statistical significance of BMI in influencing gut microbiome composition, we performed a permutational multivariate analysis of variance (PERMANOVA). The results indicated that baseline BMI accounted for a small yet statistically significant portion of the variance in microbiome composition (R² = 0.0042, p = 0.001). Furthermore, gender (R² = 0.0052, p = 0.001) and age (R² = 0.0063, p = 0.001) significantly contributed to the variation in microbial profiles. However, the majority of the variance remained unexplained (Residual R² = 0.9838), suggesting that other host and environmental factors may play a more substantial role in shaping the gut microbiome.

### Gut Microbial Taxa and Functional Pathways Associated with BMI at Baseline

The regression analysis identified several bacterial taxa that were significantly associated with BMI at baseline after FDR correction (See Table S3). Among the taxa negatively associated with BMI, Oscillospiraceae (β = -0.00983, FDR = 7.06e-08), UCG-005 (β = -0.01311, FDR = 5.28e-07), and Ruminococcaceae (β = -0.01070, FDR = 9.57e-06) exhibited the strongest inverse correlations. Other negatively associated taxa included *Faecalibacterium* (β = -0.00816, FDR = 1.46e-04), Christensenellaceae R-7 group (β = -0.00972, FDR = 5.41e-04), *Ruminococcus* (β = -0.00699, FDR = 1.50e-02) and *Akkermansia* (β = -0.00856, FDR = 2.40e-02).

Conversely, several bacterial taxa demonstrated a significant positive association with BMI. *Lachnoclostridium* (β = 0.00938, FDR = 4.51e-08), *Dorea* (β = 0.00853, FDR = 3.34e-06), *Roseburia* (β = 0.00945, FDR = 1.57e-05) and Lachnospiraceae (β = 0.00445, FDR = 1.66e-02) were among the taxa most strongly correlated with higher BMI. These findings indicate that certain taxa may be enriched in individuals with higher BMI, while others are more abundant in those with lower BMI, highlighting potential microbial contributions to obesity-related phenotypes.

Several microbial pathways were significantly associated with BMI, suggesting their potential role in obesity and metabolic health (See Table S4). Among these, the arginine degradation III pathway (mf0053, β = -0.0181, FDR = 2.42e-12) exhibited one of the strongest negative associations with BMI, highlighting its potential protective role against obesity. This pathway is involved in neurotransmitter precursor synthesis, immune modulation, and nitric oxide metabolism, indicating its role in both gut-brain communication and systemic metabolic regulation.

Other pathways associated with neurotransmitter metabolism and gut-brain axis regulation were also inversely associated with BMI. The dopamine synthesis pathway (mgb012, β = -0.0162, FDR = 5.28e-10) and serotonin synthesis I & II pathway (mgb001 & mgb002, β = -0.0155, FDR = 2.30e-06) showed negative associations, reinforcing their potential contribution to neuroendocrine signaling and appetite regulation.

With regard to the gut-liver axis, several pathways linked to amino acid metabolism and hepatic function demonstrated strong inverse associations with BMI. The tryptophan degradation pathway (mgb049, β = -0.0047, FDR = 1.22e-04) and kynurenine degradation pathway (mgb018, β = -0.0136, FDR = 2.04e-05) both negatively correlated with BMI and play roles in hepatic detoxification and systemic inflammation control. The acetate degradation pathway (mgb047, β = -0.0035, FDR = 0.0006) was inversely associated with BMI, emphasizing its role in short-chain fatty acid metabolism and hepatic energy balance. Pathways involved in inflammation and immunity also showed associations with BMI. The butyrate synthesis II pathway (mgb053, β = -0.0026, FDR = 0.0007) was inversely correlated with BMI, supporting its well-known anti-inflammatory and gut-barrier protective functions. The isovaleric acid synthesis II pathway (mgb035, β = -0.0026, FDR = 0.0003) was also negatively associated with BMI, consistent with its role in branched-chain fatty acid metabolism and gut microbial homeostasis.

Lastly, pathways influencing insulin resistance and glucose metabolism exhibited significant relationships with BMI. The acetate to acetyl-CoA pathway (mf0075, β = -0.0035, FDR = 0.0006) was negatively associated with BMI and is a key regulator of energy homeostasis and lipid metabolism.

Notably, a few microbial pathways were positively associated with BMI, suggesting a potential contribution to metabolic dysregulation. For example, the lactate production pathway (mf0092, β = 0.0014, FDR = 0.0376) and the Pentose Phosphate pathway (mf0070, β = 0.0019, FDR = 0.0266) were positively associated with BMI, indicating its possible role in inflammation modulation and gluconeogenesis. The lactose and galactose degradation pathway (mf0007, β = 0.0047, FDR = 0.0098) was also positively associated with BMI, suggesting its involvement in hepatic sugar metabolism and insulin interactions.

Taken together, these findings underscore the multifaceted roles of gut microbial pathways in obesity and metabolic regulation, with key pathways influencing the gut-brain axis, gut-liver axis, immune system, and insulin resistance.

### Polygenic Risk for BMI and Its Interaction with Gut Microbial Features

Regression analysis examined the association between PGS and BMI at baseline. While most associations were not statistically significant after FDR correction, a few PGS showed notable trends (See Table S5). As expected, the PGS for BMI itself exhibited the strongest positive association with BMI (β = 0.0194, p = 5.60E-12), reinforcing the known genetic predisposition to body weight regulation.

Further, in the regression model, we performed a post hoc analysis with the polygenic score for BMI, which was included as interaction terms with bacterial taxa and pathways that were significantly associated (FDR<0.05) with baseline BMI. The aim was to examine how a genetic predisposition to BMI modifies the relationship between gut microbiome composition and BMI. Although the interaction effects did not pass multiple testing corrections, a few notable trends were observed (Table S6). *Erysipelatoclostridium* exhibited a significant negative interaction with BMI-PGS (β = -0.274, p = 0.004), suggesting that its association with BMI is weaker in individuals with a higher genetic predisposition to obesity. Similarly, microbial pathways involved in tryptophan degradation (mgb049_tryptophan_degradation, β = -0.276, p = 0.010), lysine degradation (mf0057_lysine_degradation_i, β = -0.277, p = 0.010), and butyrate synthesis (mgb053_butyrate_synthesis_ii, β = -0.272, p = 0.012) also demonstrated negative interactions with BMI-PGS. These results indicate that in individuals with a high genetic risk for obesity, the influence of these metabolic pathways on BMI is attenuated.

In contrast, Faecalibacterium exhibited a significant positive interaction with BMI-PGS (β = 0.239, p = 0.014), implying that its association with BMI is stronger in individuals with higher genetic risk. This suggests that *Faecalibacterium*, which is typically associated with gut health and anti-inflammatory properties, may have a more pronounced impact on BMI in genetically predisposed individuals.

Although these interactions did not pass multiple testing corrections, the results suggest a potential direction of interaction between genetic risk for BMI and gut microbial features that warrant further investigation. While certain microbial functions, such as butyrate production and amino acid degradation, appear to have a diminished effect in individuals with high BMI-PGS, *Faecalibacterium* shows an enhanced association with BMI, possibly reflecting a compensatory role in these individuals. These findings highlight the need for further studies to better understand the interplay between host genetics and the gut microbiome in obesity and metabolic health.

### Associations Between Changes in Microbial Features and BMI Reduction

The regression analysis examining changes in microbial taxa and pathways (T1-T2) with the percent change in BMI was performed in a subset of individuals with gut microbiome data available at two time points (n = 180). While the associations did not remain significant after multiple testing corrections, they suggest potential trends warranting further investigation.

Among the microbial taxa that were significantly associated with baseline BMI, Christensenellaceae_R_7_group (Table S7) showed a negative association with BMI percent change (β = -0.041, p = 0.006), indicating that individuals who experienced a greater reduction in BMI tended to have an increase in this taxon over time. In contrast, *Dorea* exhibited a positive association with BMI percent change (β = 0.021, p = 0.018), suggesting that individuals with greater BMI reduction experienced a decline in this taxon. For microbial pathways that were significantly associated with baseline BMI, arginine degradation (mf0055_arginine_degradation_v) was positively associated (Table S8) with BMI percent change (β = 0.0068, p = 0.020). This suggests that individuals with greater BMI reduction had a decrease in this pathway’s activity.

## Discussion

The gut microbiome plays a key role in obesity by modulating several metabolic pathways, including the gut-brain axis, gut-liver axis, inflammation and immunity, and insulin resistance and glucose metabolism. Here, we contextualize the associations uncovered in our analysis between bacterial taxa, microbial metabolic pathways, and baseline BMI while examining how genetic risk for obesity modulates these relationships through the lens of these four key metabolic dimensions.

The gut-brain axis emerged as a significant component in the microbiome-BMI relationship, with multiple pathways related to neurotransmitter metabolism exhibiting negative associations with BMI. Pathways involved in dopamine synthesis, serotonin synthesis I and II, and tryptophan degradation were inversely correlated with BMI, suggesting a potential role of these neurotransmitters in appetite regulation and energy homeostasis. Serotonin, synthesized in the gut, plays a crucial role in satiety and gut motility, while dopamine is involved in reward-driven feeding behaviors [26]. The negative association of these pathways with BMI suggests that lower microbial contributions to neurotransmitter synthesis may be linked to increased food intake and disrupted appetite control in individuals with higher BMI. Furthermore, the interaction analysis between polygenic risk scores for BMI and gut microbial features identified a significant negative interaction for tryptophan degradation, reinforcing the potential genetic modulation of microbial contributions to neuroendocrine signaling [27].

The gut-liver axis plays a critical role in metabolic regulation and obesity, with microbial metabolism influencing hepatic function, amino acid processing, and systemic inflammation [28]. Our findings revealed a connection between BMI and microbial pathways associated with hepatic metabolism, amino acid degradation, and SCFA fermentation. Specifically, tryptophan and kynurenine degradation pathways, which are essential for hepatic detoxification, immune modulation, and neurotransmitter synthesis, were inversely associated with BMI [8,29]. This suggests that reduced microbial activity in these pathways may contribute to metabolic dysfunction, chronic inflammation, and impaired gut-brain signaling in obesity.

Emerging evidence suggests that arginine metabolism is another critical factor influencing gut-liver interactions. Arginine is a precursor for nitric oxide, which is vital in vascular function and metabolic regulation [30]. Studies indicate that alterations in arginine degradation pathways may contribute to obesity-related metabolic shifts, potentially affecting hepatic function and systemic inflammation [30,31]. Notably, L-arginine supplementation has been shown to decrease Trimethylamine N-oxide (TMAO), a microbial metabolite implicated in cardiovascular and metabolic diseases [30]. These findings align with our study’s observation that arginine degradation pathways interact with BMI dynamics, suggesting a role in obesity-related metabolic inflexibility.

The microbial metabolism of SCFA, especially butyrate and acetate, plays a central role in the gut-liver axis and energy metabolism. Butyrate, produced through microbial fermentation of fiber, is crucial in maintaining gut barrier integrity, reducing inflammation, and enhancing insulin sensitivity. Acetate degradation impacts hepatic SCFA utilization and lipid homeostasis [32,33]. Our study found that butyrate synthesis and acetate degradation were significantly associated with BMI. These associations suggest that obesity may be linked to a microbiome shift away from impaired production and hepatic SCFA utilization, thereby reducing their protective metabolic effects and influencing lipid homeostasis.

Interestingly, recent studies show how dietary fibers modulate amino acid metabolism by steering toward beneficial SCFA production [34]. These findings highlight the intricate relationship between dietary composition, gut microbial metabolism, and host metabolic health, emphasizing the need for targeted dietary interventions that enhance SCFA production to improve metabolic flexibility and obesity-related outcomes.

Chronic low-grade inflammation is a hallmark of obesity, and the gut microbiome plays a key role in immune regulation. The present study identified several microbial taxa with strong associations with BMI, particularly taxa such as Oscillospiraceae, *Faecalibacterium*, and Christensenellaceae R-7 group, which were inversely correlated with BMI. These taxa are known for their anti-inflammatory properties and have been associated with improved gut barrier integrity and metabolic health [35]. Conversely, taxa such as *Lachnoclostridium* and *Dorea*, which were positively correlated with BMI, have been linked to inflammatory conditions and gut dysbiosis [36]. Additionally, butyrate synthesis II, a key pathway for short-chain fatty acid production with well-established anti-inflammatory and gut-barrier protective effects [37,38], was negatively associated with BMI. These findings suggest that obesity is associated with an imbalance in microbial communities that regulate immune homeostasis, contributing to systemic inflammation and metabolic dysregulation.

Insulin resistance and glucose metabolism are also significantly influenced by the gut microbiome. The study identified key carbohydrate and energy metabolism pathways that exhibited associations with BMI. The acetate-to-acetyl-CoA pathway, lactate production, and lactose and galactose degradation were among the pathways linked to BMI, suggesting a role for microbial fermentation products in metabolic health [39]. Acetate and lactate serve as important metabolic intermediates that influence hepatic gluconeogenesis and lipid metabolism [40,41]. While the acetate-to-acetyl-CoA pathway was negatively associated with BMI, reinforcing its potential role in promoting metabolic efficiency, the lactate production pathway showed both positive and negative associations, suggesting a context-dependent role in obesity-related metabolic regulation. These findings indicate that microbial contributions to glucose and lipid metabolism may differentially impact obesity, with some metabolic products promoting insulin sensitivity while others may contribute to dysregulation.

The integration of genetic risk into the microbiome-BMI relationship further underscores the complex interplay between host and microbial factors in obesity. The polygenic risk score for BMI exhibited the strongest association with baseline BMI, reinforcing the known genetic contribution to body weight regulation [42]. However, interactions between BMI-PGS and microbial taxa/pathways suggest that genetic predisposition may modify the microbiome’s role in metabolic regulation. The negative interaction of *Erysipelatoclostridium* with BMI-PGS suggests that the microbial association with BMI may be attenuated in individuals with a higher genetic risk for obesity. Similarly, negative interactions observed for butyrate synthesis, tryptophan degradation, and lysine degradation pathways suggest that in genetically high-risk individuals, these microbial metabolic functions may have a reduced influence on BMI regulation. In contrast, *Faecalibacterium* exhibited a positive interaction with BMI-PGS, indicating a potentially compensatory role in genetically predisposed individuals. Although these associations lacked statistical significance after multiple test corrections, these findings highlight the need for further studies investigating the gene-microbiome interplay in obesity and metabolic health.

Lastly, we focused on understanding how microbial taxa and pathways previously associated with baseline BMI shifted following the intervention. While these associations did not pass multiple testing corrections, they reveal potential trends that warrant further exploration. Christensenellaceae, a taxon linked to leaner phenotypes and improved metabolic health [43], increased in individuals with more significant BMI reduction, suggesting a beneficial shift. Conversely, the decline in *Dorea*, a genus previously associated with obesity and metabolic dysfunction [39], indicates a possible transition toward a healthier microbiome composition. Additionally, changes in arginine metabolism, a pathway involved in nitric oxide synthesis and gut microbiome-host interactions [31,44], may reflect metabolic adaptations alongside BMI fluctuations. Given the role of nitric oxide in vascular function and metabolic regulation, these shifts could have broader implications for cardiometabolic health. Although these findings require further validation, they offer valuable insights into the dynamic relationship between gut microbiome modulation and metabolic responses to dietary and lifestyle interventions.

### Limitations of the study

Despite the strengths of this study, including a large cohort with gut microbiome and genetic data, several limitations should be acknowledged. First, the study’s observational nature limits causal inferences, and the associations identified require validation in interventional or longitudinal studies. Second, while the study identified numerous significant associations, many did not pass multiple testing corrections, indicating the need for replication in independent cohorts. Third, dietary and lifestyle factors, which are known to influence the gut microbiome, were not explicitly accounted for in the analysis, potentially introducing confounding effects. Additionally, while the longitudinal analysis provided insight into microbial changes associated with BMI reduction, the sample size for this subset was smaller (n=180), limiting statistical power. Future studies incorporating dietary, metabolic, and multi-omics data will be essential to better understand the mechanistic role of the microbiome in obesity.

## Conclusion

This study provides evidence that the gut microbiome and host genetics play a crucial role in obesity and metabolic regulation. The findings suggest that microbial taxa and pathways influence obesity through their impact on the gut-brain axis, gut-liver axis, immune regulation, and glucose metabolism. Genetic predisposition to obesity appears to modulate these relationships, potentially attenuating the microbiome’s metabolic influence in high-risk individuals. While these findings offer valuable insights, further mechanistic and interventional studies are required to fully elucidate the causal relationships between the microbiome, genetics, and obesity-related metabolic pathways. Understanding these interactions will be critical for developing personalized microbiome-based interventions for obesity and metabolic disorders.

**Table 1:** Permutational Multivariate Analysis of Variance (PERMANOVA) Assessing the Effect of BMI, Gender, and Age on Gut Microbiome Composition. Statistical significance was assessed with 1,000 permutations.

|  | Df | SumOfSqs | R<sup>2</sup> | F | Pr(>F) |
| --- | --- | --- | --- | --- | --- |
| BMI at baseline | 1 | 8210.328 | 0.004154865 | 14.28315 | 0.001 |
| Gender | 1 | 10249.400 | 0.005186745 | 17.83044 | 0.001 |
| Age | 1 | 12519.489 | 0.006335531 | 21.77961 | 0.001 |
| Residual | 3382 | 1944062.027 | 0.983799445 | NA | NA |
| Total | 3385 | 1976075.547 | 1.000000000 | NA | NA |

## Statements

### Data Availability Statement

The microbiome sequence data used in this study were submitted to NCBI BioProject ID PRJNA1245286. Supplementary materials provide individual-level data and other details, including polygenic scores, statistical results, and processed gut microbiome abundances (See Table S1-S10).

### Author Contribution

SVK, IP, DEA, and RS: Designed the study. CI, GS, RS, and DEA: Coordinated, monitored, and executed the study and data collection. SVK, IP, KMM, and SKS: collected data, processed, curated, and cleaned the data, and performed data analyses. SVK, IP, DEA, and RS wrote the manuscript. All authors reviewed and approved the manuscript.

